# Cytokine status of patients with atherosclerotic injury of carotid arteries

**DOI:** 10.1101/2021.02.19.21252052

**Authors:** M.A. Danilova, T.V. Baidina

## Abstract

The research was aimed at studying interleukin-6 (IL-6), interleukin-10 (IL-10) and C-reactive protein (CRP) levels in serum of 61 patients suffering from atherosclerotic injury of carotid arteries. The results received by the immunoferment method were compared with ultrasound features of atherosclerotic plaques and morphological peculiarities of their biopsies which were obtained during carotid endarterectomy.

An increased concentration of cytokines under analysis and CRP in serum of patients with atherosclerotic injury of carotid arteries was detected. It was found out that high levels of IL-6 and CRP are associated with progressing atherosclerosis of carotid arteries and its cerebral complications. The research revealed that detection of cytokines and CRP in serum can be used as an additional method to diagnose unstable character of atherosclerotic plaques.

## 1. Introduction

The formation of the atherosclerotic plaque in carotid arteries with further development of their stenosis or occlusion is one of the main reasons for ischemic brain injury which can cause considerable reduction in brain tissue perfusion as a result of stenosis or thrombosis and (or) embolisation of distal branches of an injured artery [13].

Atherosclerotic injury of carotid arteries in approximately 50% of patients begins with an ischemic stroke without preceding temporal disruption of blood circulation or signs of discirculatory encephalopathy [3]. Hence, diagnostics and treatment of atherosclerotic injury of carotid arteries plays a key role in prevention of an ischemic stroke. The necessity of carotid endarterectomy as a prevention method of ischemic disruption of blood flow is acknowledged in patients with stenosis of carotid arteries exceeding 70% or with presence of unstable atherosclerotic plaques in carotid arteries with stenosis more than 50%.

The efficiency of the operation in question is confirmed by a range of full-scope multi-centre investigations conducted in the USA and Europe. Active search for conservative methods of atherosclerosis treatment based on studying inflammatory mechanisms of vessel wall injury and formation of organ complications of the disease is also under way. Many aspects of carotid atherosclerosis are still under-researched and require profound studying [8,6].

In clinical practice it is of high importance to diagnose active atherosclerosis accompanied by unstable plaques formation in order to predict cardio-vascular problems. One of new approaches to their diagnostics involves analysis of inflammatory markers in blood [12,1,11,7] whose significance in cases of cerebrovascular pathology is not sufficiently explored. The present research was designed to study concentration of proinflammatory and anti-inflammatory cytokines in blood serum of patients with atherosclerotic injury of carotid arteries depending on ultrasound and pathomorphological features of atherosclerotic plaques and detect opportunities for applying inflammatory markers for diagnostics of the unstable atherosclerotic plaque in carotid arteries.

## 2. Materials and methods

84 patients were included: 61 of them had atherosclerotic plaques (the main group – 52 men and 9 women, mean age – 62 [58;69] years); 23 patients did not show signs of atherosclerotic injury of carotid arteries (the comparison group comprised 14 women and 9 men with age range from 28 to 76 years, the mean age – 48 [40;55] years).

In accordance with the requirements of biomedical ethics set by the World Medical Association of Helsinki in 2000 year all subjects included provided written informed consent for the research approved by the Ethics Committee of Perm State Academy of Medicine after expert assessment.

### 2.1. Instrumental diagnostic methods

All the patients underwent duplex scanning of the main head arteries on the ultrasound equipment Vivid −7 (USA) which allowed to assess presence and intensity of atherosclerotic injury of the vascular bed. According to ultrasound characteristics the plaques were classified as stable in 49 patients (homogenous hyperdense) and non-stable in 10 patients (homogenous hypodense and heterogenous hypo- and hyperdense).

All the patients of the main group had carotid endarterectomy with further pathomorphological studying of the atherosclerotic plaque.

Computed tomography and magnetic resonance imaging to verify the diagnosis of acute disruption of brain blood flow and to clarify its localization were carried out for patients having brain infarction in the anamnesis. Symptomatic carotid atherosclerosis was observed in 24 patients of the main group (19 patients suffered an atherothrombotic ischemic stroke, 5 patients had transitory ischemic attacks in the anamnesis). Symptomatology of patients that had suffered a stoke was characterised by the syndrome of total or partial infarction of the carotid basin. The computed tomography demonstrated territory infarction; carotid artery stenosis on the basis of duplex scanning constituted 50%. 37 patients did not reveal any clinical signs of carotid atherosclerosis.

### 2.2. Inflammatory markers in serum

Concentration of interleukin IL-6, IL −10 in blood serum was determined in all the patients before surgical treatment by means of solid-phase immunoferment research with the help of the set of reagents for immunoferment analysis A-8774 “Vector Best”. In 16 patients concentration of interleukin IL-6, IL −10 was checked repeatedly 3 months after carotid endarterectomy. All the patients of this group had multiple atherosclerosis. Hospitalization was aimed at planned surgical interference in either contralateral carotid or iliac arteries. The patients examined in dynamics did not differ in the main features from the group in general.

Concentration of C-reactive protein (CRP) in blood serum was determined in 53 patients of the main group and in all patients of the comparison group by a highly sensitive immunoferment method with the help of enzyme-linked immunosorbent assays (“BIOMERICA”, USA) before carotid endarterectomy. In 4 patients concentration of CRP was studied 3 months after the surgical treatment.

### 2.3. Pathomorphological analysis of atherosclerotic plaques

Atherosclerotic plaques were categorised as stable in 35 patients. The macroscopic picture showed their dense, without visible injuries cap, marked calcinosis. The plaque was removed as entire structure, but not in fragments. The histological analysis illustrated calcinosis in combination with fibrosis, to a lesser extent – with atheromatosis.

26 patients had unstable plaques which were characterised by macroscopic changes involving injuries of the fibrous cap of the plaque, presence of thrombus deposition on its surface, easy rupture of the plaque in an effort of its elimination. The histological picture revealed a thin fibrous cap, an increased amount of macrophages, inflammatory infiltration with neutrophils, lymphocytes, macrophages and endothelial cells, a huge atheromatous nucleus in the centre of the plaque, a decreased number of collagen fibres.

### 2.4. Sensitivity and specificity of the diagnostic methods

The comparative analysis of concentration of cytokines and CRP in blood serum as well as data of duplex scanning and results of further pathomorphological research of plaques was carried out. Sensitivity of duplex scanning methods, increased concentration of IL-6, IL-10 and CRP in blood serum (capacity to detect the disease when it is present) and specificity of tests (capacity to give a negative result when the disease is absent) in determining the character of the atherosclerotic plaque in carotid arteries were calculated [10].

### 2.5. Statistical analysis

All statistical analyses were carried out by means of the package of applied programmes STATISTICA v. 6.0. Variational and correlational analyses were used. Quantitative signs are presented in the form of the median and interquartile range. The assessment of validity of differences (*p*) between the groups of observation was applied with use of non-parametric methods of comparison based on qualitative and quantitative signs (Mann-Whitney Test), criterion χ^2^. The analysis of statistical dependencies was conducted with the help of Spearman’s rank correlation coefficient. A *p*-value of <0,05 was judged as statistically significant.

## 3. Results

### 3.1. Indicators of systemic inflammation of blood serum in carotid arteries atherosclerosis

The results of the research demonstrated connection between the atherosclerotic process in carotid arteries and indicators of systemic inflammation in blood serum. It was revealed that patients with atherosclerotic injury of carotid arteries had a higher concentration of IL-6, IL-10 and CRP in blood serum than patients that did not have atherosclerotic plaques in the carotid basin (table 1).

The data indicating dependence of concentration of IL-6, IL-10 and CRP on pathomorphological features of atherosclerotic plaques were received (table 2).

The research showed the link between IL-6 and CRP and complicated carotid atherosclerosis (table 3).

### 3.2. Dynamics of cytokine levels after carotid endarterectomy

The influence of carotid endarterectomy on concentration of IL-6 and IL-10 in blood serum was studied. The results of the analysis demonstrated that concentration of inflammatory markers after the treatment did not change considerably (table 4).

Concentration of CRP 3 months after carotid endarterectomy was determined in 4 patients. It was increased in all the 4 patients and amounted to 10,9; 3,2; 4,2 and 23,5 pg/l respectively. Interestingly, in 2 patients concentration became lower than it was before the operation, in 2 patients it rose in comparison with the initial figure.

### 3.3. Sensitivity and specificity of the diagnostic methods in determining the character of the atherosclerotic plaque in carotid arteries

To assess the validity of ultrasound research in diagnostics of atherosclerotic plaques in carotid arteries the data of duplex scanning were compared with the real state of plaques determined pathomorphologically (table 5).

As a result, sensitivity of duplex scanning towards the unstable plaque constituted 16,0%, specificity – 82,3%.

The analysis of opportunities for using the researched inflammatory markers in blood serum to diagnose unstable plaques comparing their high/ low concentration and the data of pathomorphological research of biopsies of the plaque received during carotid endarterectomy was carried out (table 6).

## 4. Discussion

Common humoural and cell reactions involved in the processes of atherosclerosis and inflammation point to their similarity. Participation of inflammatory reactions in the formation of atherosclerotic injury of carotid arteries is proved by the results of our research showing that patients with atherosclerosis of precerebral arteries have an increased concentration of IL-6, IL-10 and CRP in blood serum.

Along with participation in the initiation and progression of atherosclerosis cytokines play a certain role in destabilisation of atherosclerotic plaques in carotid arteries which is manifested by the increased concentration of IL-6, IL-10 and CRP in blood serum in patients with unstable atherosclerotic plaques in contrast to patients with stable plaques.

It agrees with the data from literature on the role of inflammation in transformation of a stable atherosclerotic plaque to an unstable one that were received in vitro and on animal models.

It is acknowledged that depletion of the fibrous cap (less than 65 mkm) and enlargement of the lipid nucleus (more than 30% of the plaque’s size) are important factors of destabilisation of the atherosclerotic plaque causing thrombotic complications. The inflammatory process can weaken the fibrous cap which enables rupture of the atherosclerotic plaque. Hence, high indicators of inflammatory markers in blood serum can serve as markers of vulnerable, rupture-prone plaques. The increased concentration of IL-6 and CRP in blood serum with manifest carotid atherosclerosis is clinical reflection of impact of these indices on features of the plaque. It is determined that the increased concentration of CRP is associated with presence of cerebral complications formed by mechanism of atherothrombosis.

According to E.Dosa’s data, removal of atherosclerotic plaques from the carotid artery leads to decrease in concentration of CRP in blood serum 6 weeks as well as 14 months after the surgical interference, mostly in patients with a high basal level of acute-phase protein.

The results of our research show that concentration of inflammatory markers in blood serum after removal of atheresclerotic plaques from carotid arteries did not alter significantly. It can be explained by the fact that dynamics of results are traced only in patients with repeated surgical interference in other vascular regions.

Atherosclerotic plaques in other vascular basins are likely to sustain inflammation and, consequently, a high level of cytokines.

It is known that presence of the unstable atherosclerotic plaque in carotid arteries influences the strategy of patient’s treatment, hence, detecting rupture-prone plaques is highly important for clinicians. According to the data from literature, the most available and informative method of diagnosing the spread, intensity and character of atherosclerotic injury is duplex scanning of vessels which allows to assess the level of stenosis, though a little less precisely in case of critical stenosis [4], the state of the wall and atherosclerotic plaque [3].

The method of duplex scanning of precerebral arteries in our research appeared to be not sufficiently sensitive in relation to diagnostics of instability of the atherosclerotic plaque in carotid arteries. Low informativeness of the method displays dependence of its results on the quality of the equipment and doctor’s qualifications.

The research showed that method of detection of IL-6 concentration in blood serum has sufficient sensitivity and specificity for determining the character of the atherosclerotic plaque. Detection of CRP concentration also appeared to be a highly sensitive test for determining instability of the atherosclerotic plaque. Detection of IL-10 concentration in blood serum turned out to be a method that rarely gives a false positive result. Consequently, detection of cytokine levels and CRP in blood serum of patients with atherosclerosis of carotid arteries is an additional objective method for determining qualitative characteristics of atherosclerotic plaques in carotid arteries.

## 5. Conclusion

The research revealed high levels of IL-6 and CRP concentration in serum of patients with pathomorphological signs of unstable atherosclerotic plaques in carotid arteries. The analysis demonstrated a diagnostic value of the method of IL-6 detection in blood serum for clarifying qualitative characteristics of atherosclerotic plaques in carotid arteries.

## Data Availability

The datasets generated during and/or analysed during the current study are available from the corresponding author on reasonable request.

